# Gender-Based Disparities in COVID-19 Patient Outcomes: A Propensity-matched Analysis

**DOI:** 10.1101/2020.04.24.20079046

**Authors:** Shailendra Singh, Monica Chowdhry, Arka Chatterjee, Ahmad Khan

## Abstract

COVID-19 epidemiological data show higher mortality among males as compared to females. However, it remains unclear if this disparity is due to gender differences in high-risk characteristics. Our study, including a large cohort of male and female patients, showed that males have a higher risk for mortality, hospitalization and mechanical ventilation even when compared to a matched cohort of females with similar age, high-risk behavior, and comorbidities. This gender-based risk of poor outcomes among COVID-19 patients is especially more pronounced in advanced age. High-risk characteristics only partially explain the gender disparity, and further research is needed to understand the causes of this disparity.

## Introduction

Coronavirus disease 2019 (COVID-19) has emerged as unprecedented global health, economic, and social crisis. More than 2 million COVID-19 cases and 160,00 deaths have been reported worldwide as of April 20, 2020^1,2^. Epidemiological data reported across multiple countries show markedly higher mortality for males compared to females^3,4^. It needs to be determined if the gender disparity in the COVID-19 mortality is true or just an interaction of gender differences in risk characteristics^5^. Therefore, we studied the clinical characteristics of a large and diverse cohort of COVID-19 patients stratified by gender and determined the outcomes after extensive propensity matching for risk factors and comorbidities.

## Methods

The study was conducted using TriNetX (Cambridge, MA, USA), a global federated health research network providing real-time access to electronic medical records (EMR) of more than 49 million patients from 37 healthcare organizations (HCOs) predominantly in the United States. COVID-19 Research Network was recently created, data inflow fast-tracked with COVID-19 specific diagnosis and terminology following the WHO and CDC COVID-19 guidelines^6–8^. As a federated network, TriNetX received a waiver from Western IRB since only aggregated counts, statistical summaries of de-identified information, but no protected health information is received, and no study-specific activities are performed in retrospective analyses. Both the patients and HCO’s as data sources stay anonymous.

A search query was made (April 17-18, 2020) in the EMR of patients on the COVID-19 research network to identify male and female patients (≥ 10 years age) diagnosed with COVID-19 between January 20, 2020, and April 15, 2020. The primary outcome was risk of mortality, mechanical ventilation, and hospitalization within 30 days after the diagnosis of COVID-19.

All statistical analyses were performed in real-time using TriNetX. TriNetX obfuscates patient counts to safeguard protected health information (PHI) by rounding patient counts in analyses up to the nearest 10. A 1:1 propensity score matching was performed using a greedy nearest-neighbor matching algorithm. For each outcome, the risk difference and risk ratio were calculated. We also performed Kaplan-Meier survival analyses to estimate the survival probability of each outcome at the end 30 days after the diagnosis of COVID-19. A-priori defined two-sided alpha of less than <0.05 was used for statistical significance.

Details of data source, search criteria, the definition of variables, and statistical analysis are available in Supplementary.

## Results

We identified a total of 5980 males and 7730 females diagnosed with COVID-19 from 20 HCOs. Of these, 45% of males and 50% of females were from the United States. Males were older than the females (54.9 ± 18.3 vs. 50.9 ± 18.4, p-value <0.0001). There were significant differences in patient demographics, risk behaviors, and comorbidities among male and female patients with COVID-19 (Table 1). Age, race, high-risk behavior including nicotine, and all comorbidities (mentioned in Table 1), including diabetes, hypertension, chronic lung disease, cardiovascular disease, chronic kidney disease, among others, were selected for propensity matching. The groups (N=5350 each group) were well balanced after matching (Table 1). Presenting signs and symptoms and most recent laboratory findings after COVID-19 are also compared in Table 1. Mean values of serum creatinine, liver function tests, and inflammatory markers such as Ferritin, CRP, and IL-6 were all significantly higher in males.

**Table 1:**
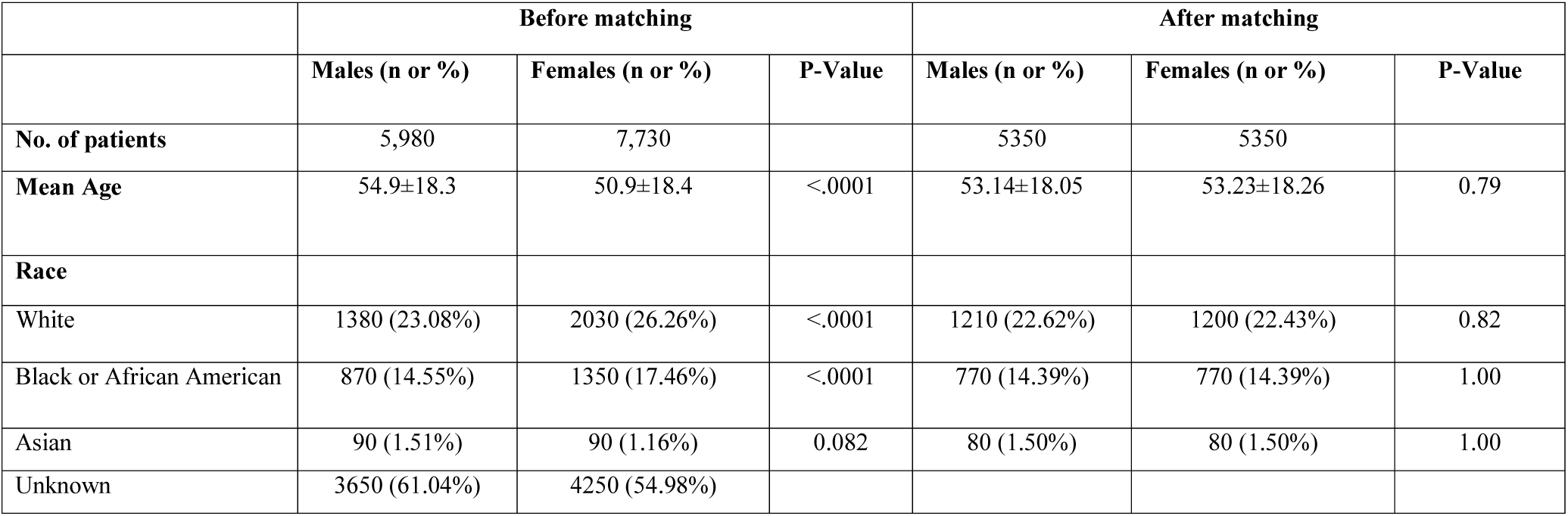
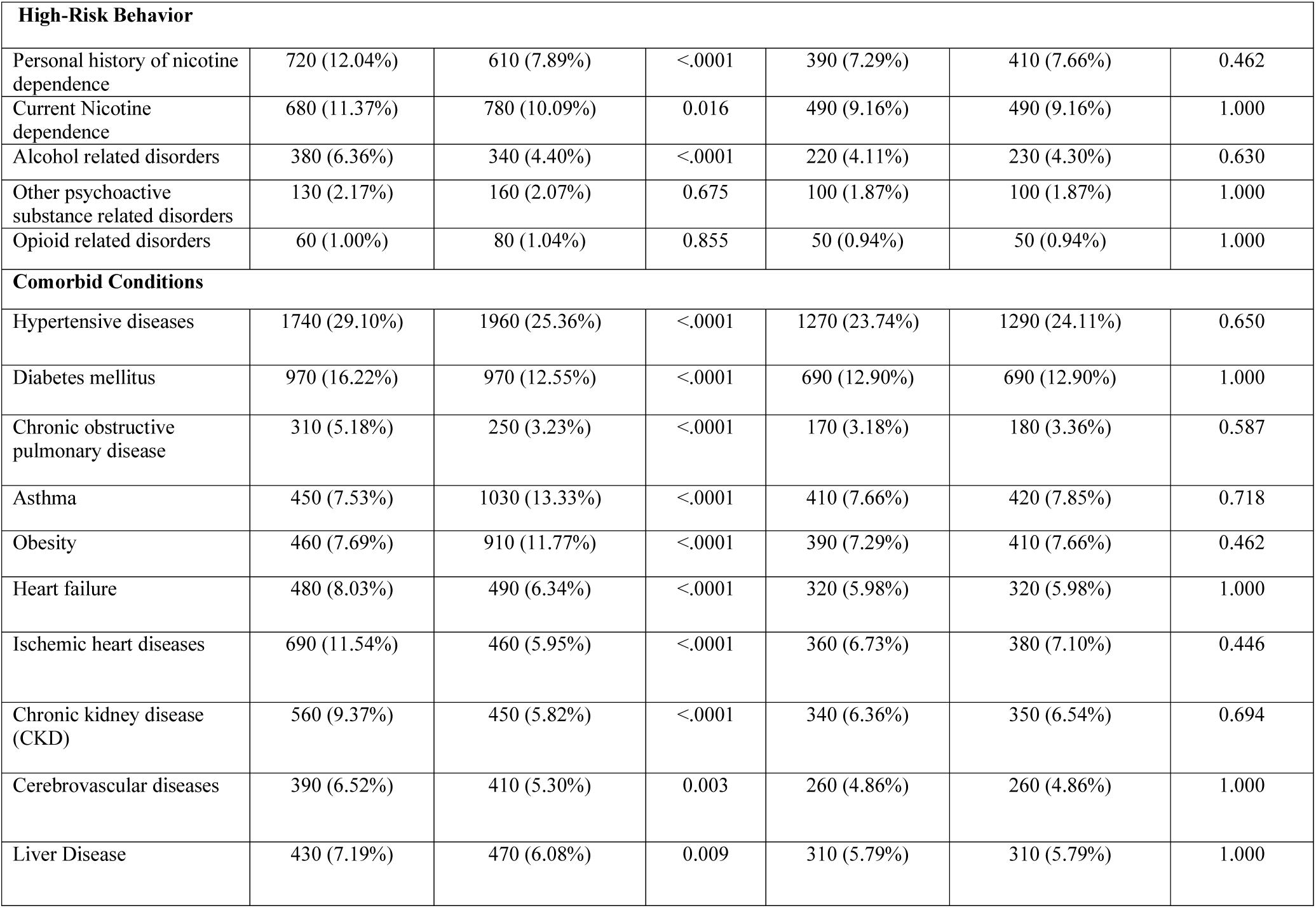
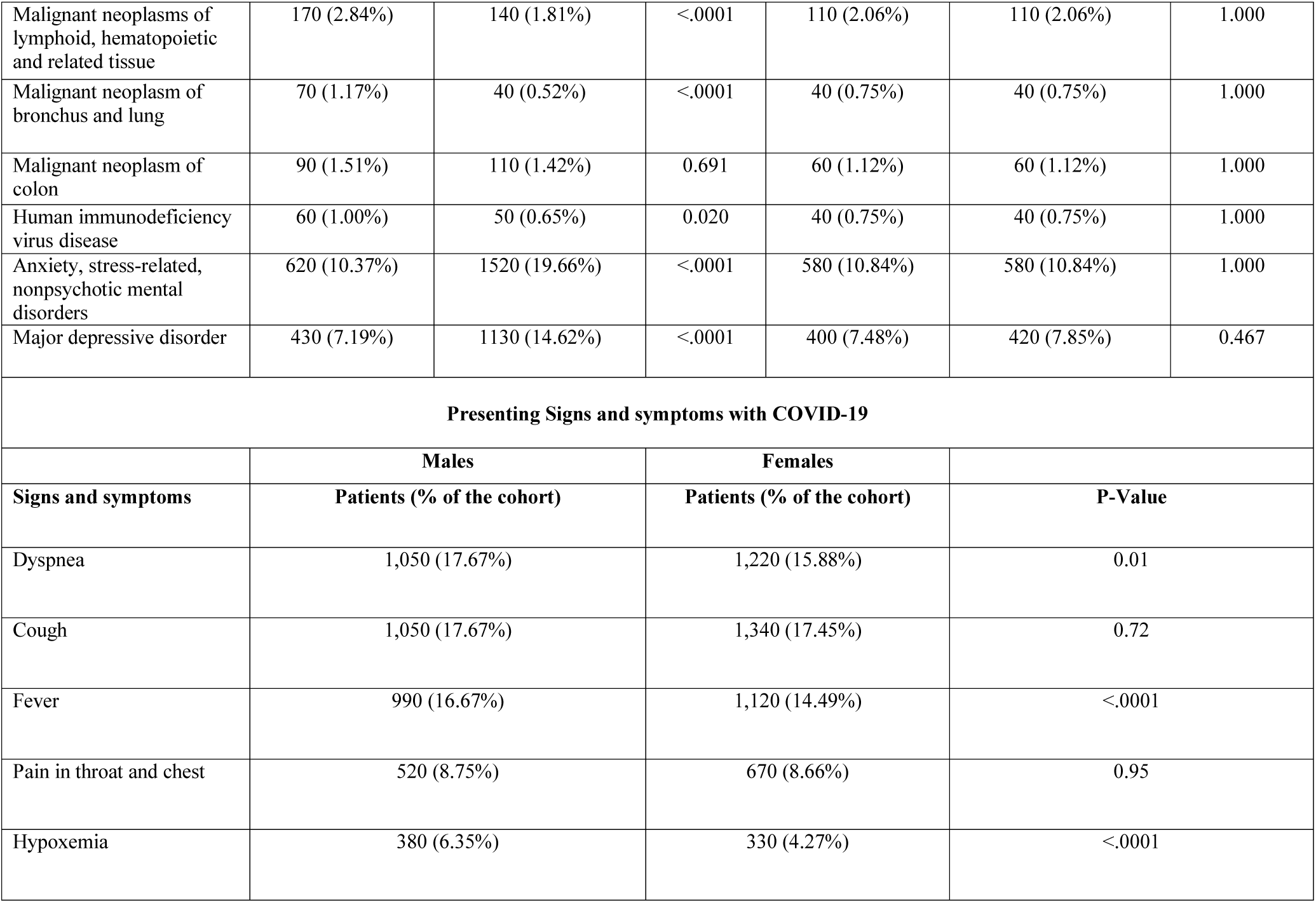
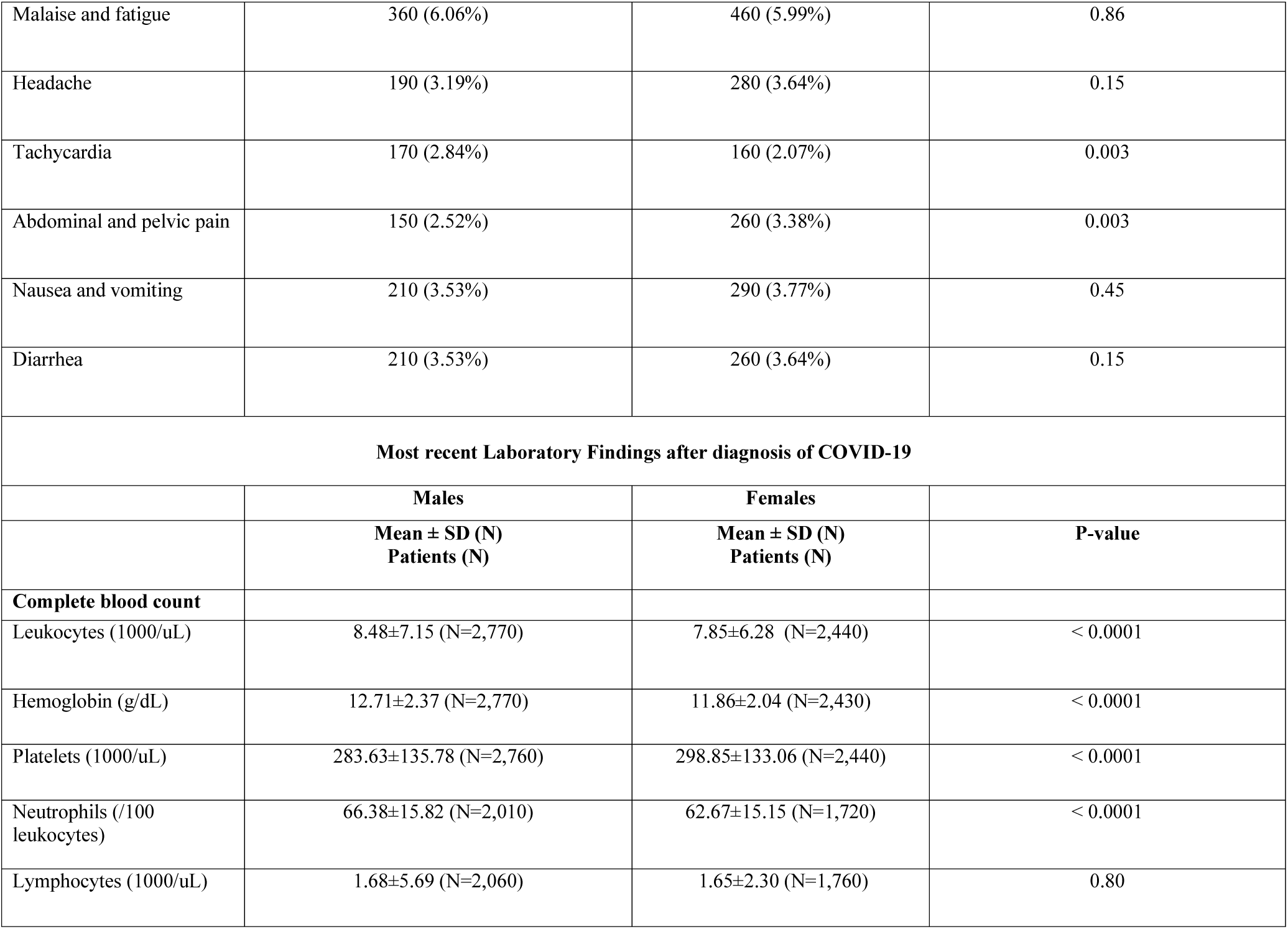
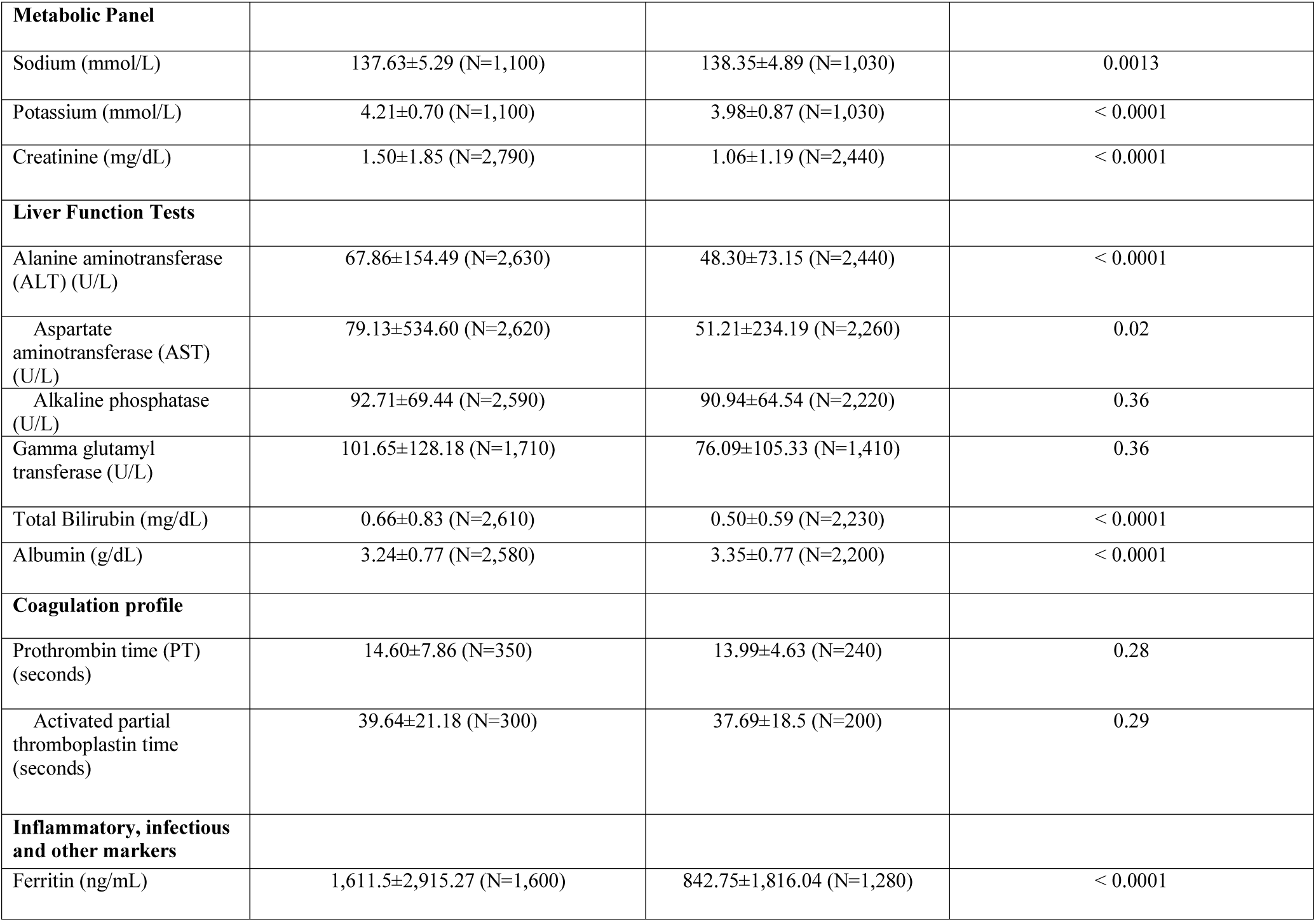
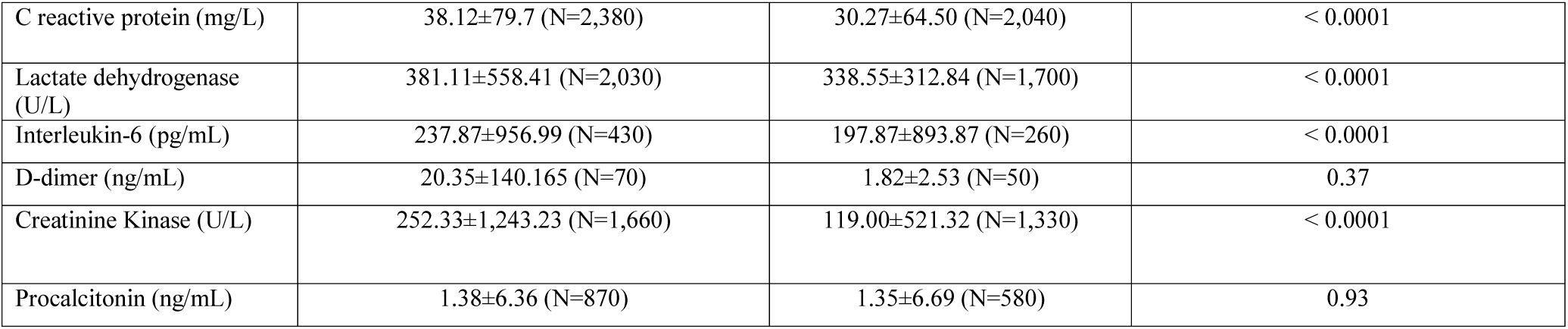
Comparison of patient characteristics, presenting signs and symptoms, and laboratory findings among COVID-19 male and female patients. Demographics and comorbidities are compared before and after propensity matching of cohorts.

|  | Before matching |  |  | After matching |  |  |
| --- | --- | --- | --- | --- | --- | --- |
|  | Males (n or %) | Females (n or %) | P-Value | Males (n or %) | Females (n or %) | P-Value |
| **No. of patients** | 5,980 | 7,730 |  | 5350 | 5350 |  |
| **Mean Age** | 54.9±18.3 | 50.9±18.4 | <.0001 | 53.14±18.05 | 53.23±18.26 | 0.79 |
| **Race** |  |  |  |  |  |  |
| White | 1380 (23.08%) | 2030 (26.26%) | <.0001 | 1210 (22.62%) | 1200 (22.43%) | 0.82 |
| Black or African American | 870 (14.55%) | 1350 (17.46%) | <.0001 | 770 (14.39%) | 770 (14.39%) | 1.00 |
| Asian | 90 (1.51%) | 90 (1.16%) | 0.082 | 80 (1.50%) | 80 (1.50%) | 1.00 |
| Unknown | 3650 (61.04%) | 4250 (54.98%) |  |  |  |  |
| **High-Risk Behavior** |  |  |  |  |  |  |
| Personal history of nicotine dependence | 720 (12.04%) | 610 (7.89%) | <.0001 | 390 (7.29%) | 410 (7.66%) | 0.462 |
| Current Nicotine dependence | 680 (11.37%) | 780 (10.09%) | 0.016 | 490 (9.16%) | 490 (9.16%) | 1.000 |
| Alcohol related disorders | 380 (6.36%) | 340 (4.40%) | <.0001 | 220 (4.11%) | 230 (4.30%) | 0.630 |
| Other psychoactive substance related disorders | 130 (2.17%) | 160 (2.07%) | 0.675 | 100 (1.87%) | 100 (1.87%) | 1.000 |
| Opioid related disorders | 60 (1.00%) | 80 (1.04%) | 0.855 | 50 (0.94%) | 50 (0.94%) | 1.000 |
| **Comorbid Conditions** |  |  |  |  |  |  |
| Hypertensive diseases | 1740 (29.10%) | 1960 (25.36%) | <.0001 | 1270 (23.74%) | 1290 (24.11%) | 0.650 |
| Diabetes mellitus | 970 (16.22%) | 970 (12.55%) | <.0001 | 690 (12.90%) | 690 (12.90%) | 1.000 |
| Chronic obstructive pulmonary disease | 310 (5.18%) | 250 (3.23%) | <.0001 | 170 (3.18%) | 180 (3.36%) | 0.587 |
| Asthma | 450 (7.53%) | 1030 (13.33%) | <.0001 | 410 (7.66%) | 420 (7.85%) | 0.718 |
| Obesity | 460 (7.69%) | 910 (11.77%) | <.0001 | 390 (7.29%) | 410 (7.66%) | 0.462 |
| Heart failure | 480 (8.03%) | 490 (6.34%) | <.0001 | 320 (5.98%) | 320 (5.98%) | 1.000 |
| Ischemic heart diseases | 690 (11.54%) | 460 (5.95%) | <.0001 | 360 (6.73%) | 380 (7.10%) | 0.446 |
| Chronic kidney disease (CKD) | 560 (9.37%) | 450 (5.82%) | <.0001 | 340 (6.36%) | 350 (6.54%) | 0.694 |
| Cerebrovascular diseases | 390 (6.52%) | 410 (5.30%) | 0.003 | 260 (4.86%) | 260 (4.86%) | 1.000 |
| Liver Disease | 430 (7.19%) | 470 (6.08%) | 0.009 | 310 (5.79%) | 310 (5.79%) | 1.000 |
| Malignant neoplasms of lymphoid, hematopoietic and related tissue | 170 (2.84%) | 140 (1.81%) | <.0001 | 110 (2.06%) | 110 (2.06%) | 1.000 |
| Malignant neoplasm of bronchus and lung | 70 (1.17%) | 40 (0.52%) | <.0001 | 40 (0.75%) | 40 (0.75%) | 1.000 |
| Malignant neoplasm of colon | 90 (1.51%) | 110 (1.42%) | 0.691 | 60 (1.12%) | 60 (1.12%) | 1.000 |
| Human immunodeficiency virus disease | 60 (1.00%) | 50 (0.65%) | 0.020 | 40 (0.75%) | 40 (0.75%) | 1.000 |
| Anxiety, stress-related, nonpsychotic mental disorders | 620 (10.37%) | 1520 (19.66%) | <.0001 | 580 (10.84%) | 580 (10.84%) | 1.000 |
| Major depressive disorder | 430 (7.19%) | 1130 (14.62%) | <.0001 | 400 (7.48%) | 420 (7.85%) | 0.467 |
| **Presenting Signs and symptoms with COVID-19** |  |  |  |  |  |  |
|  | **Males** |  | **Females** |  |  |  |
| **Signs and symptoms** | **Patients (% of the cohort)** |  | **Patients (% of the cohort)** |  | **P-Value** |  |
| Dyspnea | 1,050 (17.67%) |  | 1,220 (15.88%) |  | 0.01 |  |
| Cough | 1,050 (17.67%) |  | 1,340 (17.45%) |  | 0.72 |  |
| Fever | 990 (16.67%) |  | 1,120 (14.49%) |  | <.0001 |  |
| Pain in throat and chest | 520 (8.75%) |  | 670 (8.66%) |  | 0.95 |  |
| Hypoxemia | 380 (6.35%) |  | 330 (4.27%) |  | <.0001 |  |

|  |  |  |  |
| --- | --- | --- | --- |
| Malaise and fatigue | 360 (6.06%) | 460 (5.99%) | 0.86 |
| Headache | 190 (3.19%) | 280 (3.64%) | 0.15 |
| Tachycardia | 170 (2.84%) | 160 (2.07%) | 0.003 |
| Abdominal and pelvic pain | 150 (2.52%) | 260 (3.38%) | 0.003 |
| Nausea and vomiting | 210 (3.53%) | 290 (3.77%) | 0.45 |
| Diarrhea | 210 (3.53%) | 260 (3.64%) | 0.15 |
| **Most recent Laboratory Findings after diagnosis of COVID-19** |  |  |  |
|  | **Males** | **Females** |  |
|  | **Mean ± SD (N)**<br>**Patients (N)** | **Mean ± SD (N)**<br>**Patients (N)** | **P-value** |
| **Complete blood count** |  |  |  |
| Leukocytes (1000/uL) | 8.48±7.15 (N=2,770) | 7.85±6.28 (N=2,440) | < 0.0001 |
| Hemoglobin (g/dL) | 12.71±2.37 (N=2,770) | 11.86±2.04 (N=2,430) | < 0.0001 |
| Platelets (1000/uL) | 283.63±135.78 (N=2,760) | 298.85±133.06 (N=2,440) | < 0.0001 |
| Neutrophils (/100 leukocytes) | 66.38±15.82 (N=2,010) | 62.67±15.15 (N=1,720) | < 0.0001 |
| Lymphocytes (1000/uL) | 1.68±5.69 (N=2,060) | 1.65±2.30 (N=1,760) | 0.80 |
| **Metabolic Panel** |  |  |  |
| Sodium (mmol/L) | 137.63±5.29 (N=1,100) | 138.35±4.89 (N=1,030) | 0.0013 |
| Potassium (mmol/L) | 4.21±0.70 (N=1,100) | 3.98±0.87 (N=1,030) | < 0.0001 |
| Creatinine (mg/dL) | 1.50±1.85 (N=2,790) | 1.06±1.19 (N=2,440) | < 0.0001 |
| **Liver Function Tests** |  |  |  |
| Alanine aminotransferase (ALT) (U/L) | 67.86±154.49 (N=2,630) | 48.30±73.15 (N=2,440) | < 0.0001 |
| Aspartate aminotransferase (AST) (U/L) | 79.13±534.60 (N=2,620) | 51.21±234.19 (N=2,260) | 0.02 |
| Alkaline phosphatase (U/L) | 92.71±69.44 (N=2,590) | 90.94±64.54 (N=2,220) | 0.36 |
| Gamma glutamyl transferase (U/L) | 101.65±128.18 (N=1,710) | 76.09±105.33 (N=1,410) | 0.36 |
| Total Bilirubin (mg/dL) | 0.66±0.83 (N=2,610) | 0.50±0.59 (N=2,230) | < 0.0001 |
| Albumin (g/dL) | 3.24±0.77 (N=2,580) | 3.35±0.77 (N=2,200) | < 0.0001 |
| **Coagulation profile** |  |  |  |
| Prothrombin time (PT) (seconds) | 14.60±7.86 (N=350) | 13.99±4.63 (N=240) | 0.28 |
| Activated partial thromboplastin time (seconds) | 39.64±21.18 (N=300) | 37.69±18.5 (N=200) | 0.29 |
| **Inflammatory, infectious and other markers** |  |  |  |
| Ferritin (ng/mL) | 1,611.5±2,915.27 (N=1,600) | 842.75±1,816.04 (N=1,280) | < 0.0001 |
| C reactive protein (mg/L) | 38.12±79.7 (N=2,380) | 30.27±64.50 (N=2,040) | < 0.0001 |
| Lactate dehydrogenase (U/L) | 381.11±558.41 (N=2,030) | 338.55±312.84 (N=1,700) | < 0.0001 |
| Interleukin-6 (pg/mL) | 237.87±956.99 (N=430) | 197.87±893.87 (N=260) | < 0.0001 |
| D-dimer (ng/mL) | 20.35±140.165 (N=70) | 1.82±2.53 (N=50) | 0.37 |
| Creatinine Kinase (U/L) | 252.33±1,243.23 (N=1,660) | 119.00±521.32 (N=1,330) | < 0.0001 |
| Procalcitonin (ng/mL) | 1.38±6.36 (N=870) | 1.35±6.69 (N=580) | 0.93 |

Males had a significantly higher risk for mortality before (Risk Ratio (RR) 2.1, 95% CI 1.8-2.4) and after matching (RR 1.4, 95% CI 1.2-1.7) (Table 2). Kaplan-Meier survival curves showed significantly different 30-day survival probability for mortality after diagnosis of COVID-19 in matched male (80%) and female cohorts (87%) (p-value <0.0001) (Figure 1). The risk of hospitalization (RR 1.3, 95%CI 1.2-1.4) and mechanical ventilation (RR 1.71, 95% CI 1.3-2.3) was significantly higher in males (Table 2) even after matching of cohorts. A sub-analysis of the cohorts based on age was also performed. Patients (≥ 50 years (mean age of cohort) were considered as advanced age, and 93.9% of overall mortality was in patients ≥ 50 years. Males with advanced age had higher mortality than matched females of similar age group (RR 1.6, 95% CI 1.4-1.8) (Table 2).

**Table 2:** Outcomes among COVID-19 male and female patients. Outcomes are compared before and after propensity score matching of cohorts.

|  | Males |  | Females |  | Measure of Association |  |  |
| --- | --- | --- | --- | --- | --- | --- | --- |
| Outcomes | Patients with outcomes | Risk (%) | Patients with outcomes | Risk (%) | Risk Ratio (95% confidence interval) | Risk difference (95% confidence interval) | P-Value |
| **Before propensity score matching** |  |  |  |  |  |  |  |
| Mortality | 530/5,980 | 8.86% | 330/7,730 | 4.27% | 2.08 (1.82,2.37) | 4.59% (3.74%, 5.44%) | <0.0001 |
| Hospitalization | 2,150/5,980 | 35.95% | 1,840/7,730 | 23.80% | 1.51(1.43,1.59) | 12.15% (10.60%, 13.69%) | <0.0001 |
| Mechanical Ventilation | 140/5,980 | 2.34% | 100/7,730 | 1.29% | 1.81(1.40,2.33) | 1.05% (0.59%, 1.51%) | <0.0001 |

| **After propensity score matching** |  |  |  |  |  |  |  |
| --- | --- | --- | --- | --- | --- | --- | --- |
| Mortality | 390/5,350 | 7.29% | 270/5,350 | 5.05% | 1.44 (1.24,1.67) | 2.24% (1.33%, 3.15%) | <0.0001 |
| Hospitalization | 1,810/5,350 | 33.83% | 1,410/5,350 | 26.35% | 1.28 (1.21,1.36) | 7.48% (5.74%, 9.20%) | <0.0001 |
| Mechanical Ventilation | 120/5,350 | 2.24% | 70/5,350 | 1.31% | 1.71 (1.28,2.30) | 0.93% (0.43%, 1.43%) | 0.0003 |
| **Subgroup Age ≥ 50 years After propensity score matching** |  |  |  |  |  |  |  |
| Mortality | 380/3,020 | 12.58% | 240/3020 | 7.95% | 1.58 (1.35-1.84) | 4.63% (3.10%, 6.16%) | <0.0001 |
| Mechanical Ventilation | 80/3,020 | 2.64% | 60/3020 | 1.98% | 1.33 (0.96-1.86) | 0.66% (-0.097%, 1.42%) | 0.0872 |
| Hospitalization | 1,340/3,020 | 44.37% | 1,100/3020 | 36.42% | 1.21 (1.14-1.29) | 7.94% (5.48%, 10.41%) | <0.0001 |

**Figure 1:**
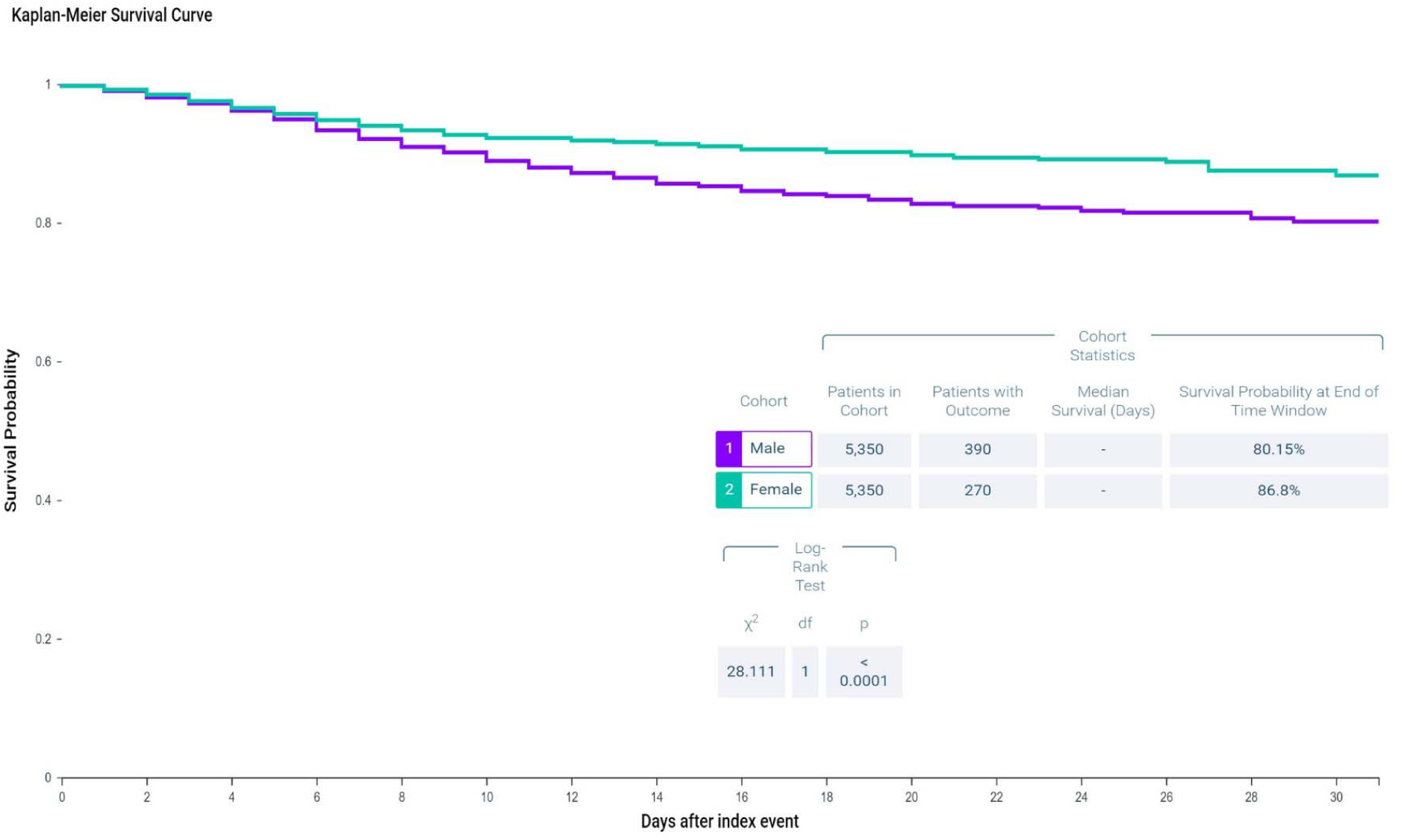
Kaplan-Meier survival curves estimating the 30-day survival probability among propensity-matched male and female cohort of COVID-19 patients.

## Discussion

Our study, including a large cohort of male and female COVID 19 patients, showed that the overall risk for mortality, hospitalization, and mechanical ventilation was significantly higher for males as compared to females. We meticulously compared the characteristics of male and female patients with COVID-19 and found a higher rate of high-risk behaviors and comorbidities in male patients. Even after propensity score matching of the cohorts for age, race, nicotine use, diabetes, hypertension, chronic lung diseases, cardiovascular diseases, chronic kidney disease, and many other comorbidities/risk behaviors, gender disparity in outcomes persist.

The vast majority of the deaths in COVID-19 occurred in patients with advanced age (≥50 years), and the gender-dependent risk of poor outcomes after COVID-19 was more pronounced with advancing age. The risk of mortality in patients with advanced age was higher compared to a matched group of females of similar age. In younger patients, the overall mortality is low, and therefore mortality differences might not be apparent. We observed higher levels of serum creatinine, liver function tests, and inflammatory markers like ferritin, CRP, and IL-6 in males, likely suggesting more severe disease.

COVID-19 gender differences in mortality are similar to the 2002-2003 SARS-CoV epidemic and the later MERS-CoV outbreak^9,10^. Multiple factors likely contribute to the disparity in gender-specific outcomes. Age and high-risk comorbid conditions could only partially explain the stark differences reported with unadjusted data. The higher smoking rate in men is speculated as a possible reason^11^, we matched our cohorts for both past and current nicotine use and still found significant differences in outcome. Moreover, the difference in rates of smoking is unlikely to explain the magnitude of gender disparity seen in outcomes. Although gender-specific variations in angiotensin-converting enzyme 2 (ACE2) receptor expression have been suggested (preprint)^12^, this needs further confirmation. Immune-defense factors likely have a substantial role. Males and females respond differently to many virus infections, and generally, females mount a more robust protective immune response^13,14^. Differences in the production of sex-specific hormones, immune response X-linked genes, and the disease-susceptibility genes among males and females likely impact the immune response and disease severity^13,14^. Besides biological factors, many social, behavioral, and cultural differences might also have a role.

Although our data represent one of the largest and most diverse cohorts of COVID-19 patients, our study has several limitations. The data derived from EMR based database is susceptible to errors in coding or data entry^15^. However, the ability of TriNetX to aggregate the data directly from the EMRs in a real-time fashion minimizes the risk of data collection errors at the investigator’s end. Patient counts were rounded up to the nearest ten, and that may influence results for infrequent outcomes; however, most of our outcomes had a relatively large number of patients. Gender differences in asymptomatic or mild cases of COVID-19 that remain undiagnosed or did not seek medical care were not accounted for^16^, and hence their role might be undervalued.

In conclusion, males are more severely affected and have higher mortality from COVID-19. This gender-specific risk is especially more pronounced in advanced age. Gender disparity in poor outcomes can only be partially explained by differences in high-risk behavior and comorbidities. Further research is needed to understand the causes of this disparity and may be beneficial for decisions in both clinical and policy-making realms.

## Data Availability

None

## Acknowledgment

We acknowledge the West Virginia Clinical and Translational Science Institute to provide us access, and training to the TriNETX global healthcare network.

We also acknowledge the TriNETX (Cambridge, MA, USA) healthcare network for design assistance to complete this project.

## References

1. COVID-19 Dashboard by the Center for Systems Science and Engineering (CSSE) at Johns Hopkins University (JHU). https://coronavirus.jhu.edu/map.html. Accessed April 19, 2020.

2. WHO Coronavirus Disease 2019 (COVID-19) Situation Report—91. https://www.who.int/docs/default-source/coronaviruse/situation-reports/20200420-sitrep-91-covid-19.pdf?sfvrsn=fcf0670b_4.

3. Team TNCPERE. The Epidemiological Characteristics of an Outbreak of 2019 Novel Coronavirus Diseases (COVID-19) — China, 2020. China CDC Wkly. 2020.

4. COVID-19 sex-disaggregated data tracker, https://globalhealth5050.org/covidl9/.

5. Liu S, Zhang M, Yang L, et al. Prevalence and patterns of tobacco smoking among Chinese adult men and women: Findings of the 2010 national smoking survey. J Epidemiol Community Health. 2017. doi:10.1136/jech-2016-207805

6. TriNetX. https://www.trinetx.com/covid-19/.

7. ICD-10-CM Official Coding Guidelines - Supplement Coding encounters related to COVID-19 Coronavirus Outbreak. https://www.cdc.gov/nchs/data/icd/ICD-10-CM-Official-Coding-Gudance-lnterim-Advice-coronavirus-feb-20-2020.pdf.

8. ICD-10-CM Official Coding and Reporting Guidelines April 1, 2020 through September 30, 2020. https://www.cdc.gov/nchs/data/icd/COVID-19-guidelines-final.pdf.

9. Karlberg J, Chong DSY, Lai WYY. Do Men Have a Higher Case Fatality Rate of Severe Acute Respiratory Syndrome than Women Do? Am J Epidemiol. 2004. doi:10.1093/aje/kwh056

10. Alghamdi IG, Hussain II, Almalki SS, Alghamdi MS, Alghamdi MM, El-Sheemy MA. The pattern of Middle east respiratory syndrome coronavirus in Saudi Arabia: A descriptive epidemiological analysis of data from the Saudi Ministry of Health. Int J Gen Med. 2014. doi:10.2147/IJGM.S67061

11. Cai H. Sex difference and smoking predisposition in patients with COVID-19. Lancet Respir Med. 2020. doi:10.1016/S2213-2600(20)30117-X

12. Zhao Y, Zhao Z, Wang Y, Zhou Y, Ma Y, Zuo W. Single-cell RNA expression profiling of ACE2, the putative receptor of Wuhan 2019-nCov. *bioRxiv*. January 2020:2020.01.26.919985. doi:10.1101/2020.01.26.919985

13. Klein SL, Flanagan KL. Sex differences in immune responses. Nat Rev Immunol. 2016. doi:10.1038/nri.2016.90

14. Channappanavar R, Fett C, Mack M, Ten Eyck PP, Meyerholz DK, Perlman S. Sex-Based Differences in Susceptibility to Severe Acute Respiratory Syndrome Coronavirus Infection. J Immunol. 2017. doi:10.4049/jimmunol.1601896

15. Bowman S. Impact of electronic health record systems on information integrity: quality and safety implications. Perspect Health Inf Manag. 2013.

16. Nishiura H, Kobayashi T, Suzuki A, et al. Estimation of the asymptomatic ratio of novel coronavirus infections (COVID-19). Int J Infect Dis. 2020. doi:10.1016/j.ijid.2020.03.020

